# Abdominal myosteatosis is associated with lower processing speed in a multiethnic cohort of older adults

**DOI:** 10.1101/2025.01.12.25320415

**Authors:** Brendan L. McNeish, Iva Miljkovic, Matthew A. Allison, Timothy Hughes, Ilya Nasrallah, Eric Terkpertey, Caterina Rosano

## Abstract

**Background:** Prior research linking myosteatosis with cognition in older adults has been conducted in relatively homogenous populations with narrow age ranges. We evaluated if abdominal myosteatosis was associated with processing speed in a multiethnic cohort of middle aged and older adults.

**Methods:** The analytical sample included 1,268 adults (46-86 years old, mean 63±9 years, 53% female of 41% White, 20% Black, 14% Chinese, and 25% Hispanic), a subset from the Multi-Ethnic Study of Atherosclerosis (MESA). Bivariate analyses were performed between abdominal computed tomography derived muscle densities (a myosteatosis measure) at year 3 with Digit Symbol Coding (DSC) with cytokines. Multivariable models were first adjusted for demographics, education, and general cognition, and further adjusted for other known predictors of dementia: *APOE*-4, physical activity, diabetes, cholesterol, smoking, and blood pressure. We further assessed whether central adiposity, general adiposity, and cytokines modified this association. We tested interactions by ethnicity, sex, and age.

**Results:** Rectus abdominis myosteatosis was significantly associated with worse DSC (B= −0.247, 95% CI: 0.098,0.396, p=0.001) independent of demographics, education, general cognition, and dementia risk factors. Adjustment for central adiposity, and cytokines did not attenuate the associations. Tests for interactions by ethnicity, sex, and age were not statistically significant.

**Conclusions:** Rectus abdominis myosteatosis is associated with worse processing speed in this middle and older aged multiethnic population of men and women, independent of other known predictors of cognition. Longitudinal studies should assess the interplay of myosteatosis with other markers of adiposity, inflammation, and circulating mediators and their combined impact on processing speed.

**Highlights:**

- Abdominal myosteatosis correlated with lower cognitive processing speed in older adults.
- Myosteatosis links muscle density to cognitive function beyond dementia risk factors.
- Pro-inflammatory cytokines do not mediate the muscle-cognition association.
- Ethnicity, sex, and adiposity measures do not modify myosteatosis-cognition links.
- Abdominal CT scans could serve as diagnostic tools for cognitive health insights

## 1. INTRODUCTION

Alzheimer’s disease (AD) and dementia contribute to the loss of mobility and independence and lead to increased mortality.^1^ Muscle health and central adiposity are emerging risk factors for declining cognitive function across populations,^2–5^ but they have rarely been studied together. Interestingly, these factors may intersect through adiposity within muscle, known as myosteatosis, which increases with age and has been associated with declines in muscle and physical function, as well as cardio-metabolic disorders.^6–10^ There are preliminary reports that increased myosteatosis is associated with greater cognitive decline in older adults.^11,12^

We have recently shown in a large cohort of Black and White adults aged 69-79 years that increases in thigh myosteatosis predicted a decline in mini mental status scores.^11^ Additionally, prior work demonstrates that higher levels of abdominal myosteatosis are associated with reduced information processing speed measured by Digit Symbol Substitution Test (DSST) in African Caribbean women younger than age 65.^12^ Moreover, two cross-sectional studies in older Caucasian European adults demonstrated a negative association between skeletal myosteatosis and neuroimaging correlates of brain aging.^13,14^

Compared to their age matched peers, Black and Hispanic adults, especially women, are disproportionally affected by AD^15–17^, while levels of myosteatosis differ based upon sex and ethnic background.^18,19^ Despite the emerging evidence for the association of increased levels of myosteatosis with lower levels of cognitive health, there is a current gap in the literature as to whether this relationship remains significant in adults from diverse racial and ethnic backgrounds and among larger age-ranges.^17^ Therefore, identifying and understanding myosteatosis’s relationship to cognitive function could provide novel scientific and clinical insights for developing targeted prevention and intervention strategies in populations at higher risk for AD, ultimately promoting more personalized and equitable healthcare.

Different fat depots in the body may not have equivalent impacts on health outcomes, particularly when it comes to cognitive function and metabolic health. For example, visceral adiposity and intramuscular fat (myosteatosis) each have unique physiological effects and may influence cardio-metabolic and inflammatory processes differently. Understanding how each type of adiposity relates to cognitive health is essential, as these fat depots may drive distinct inflammatory and metabolic pathways that alter muscle function and, consequently, cognitive outcomes. Therefore, evaluating the role of adiposity, cardio-metabolic health, and inflammatory cytokines in the relationship between muscle health and cognitive function could provide crucial insights into how specific fat deposits may modulate these associations, potentially revealing novel intervention points for rehabilitating cognitive health in at-risk populations.

The primary aim of our study was to evaluate the association between abdominal skeletal myosteatosis and processing speed in a racially and ethnically diverse cohort of middle-aged and older adults, and to determine whether this relationship remained consistent across ethnic groups, independent of known dementia risk factors, measures of general or central adiposity, and cytokine levels. Given the sex, race, and ethnicity-specific differences in skeletal muscle adiposity and dementia risk, we also tested the interaction of skeletal myosteatosis by sex, race, and ethnicity to detect potential group differences. We hypothesized that abdominal muscle density would be positively associated with processing speed, independent of dementia risk factors, central adiposity measures, and cytokine levels, and that there would be a significant interaction within Black and Hispanic older adults, but not by sex.

## 2. METHODS

### 2.1 Population

The Multi-Ethnic Study of Atherosclerosis (MESA) is a longitudinal study of adults 45-84 years of age that were recruited from six geographic regions across the United States. Details regarding the study design having been previously described.^20^ Briefly, the cohort included a total of 6,814 men and women who were without clinically apparent cardiovascular disease at the time of enrollment (July 2000 to August 2002). Between 2002 and 2005, 1,970 participants were randomly enrolled in an ancillary study during clinic visits 2 and 3, with abdominal CT scans obtained. Roughly half underwent scans at visit 2, and the remainder at visit 3. To ensure contemporaneous measurements, demographic, biomarker, and physical activity data from the corresponding visit were used. The MESA studies were approved by the institutional review board and all participants gave written informed consent.

### 2.2 Primary Independent Variable: Abdominal Muscle Measurements

Abdominal muscle, visceral fat, and intermuscular fat were measured from CT scans obtained at visits 2 or 3 and have been previously reported.^9^ Briefly, Six transverse slices were analyzed using MIPAV software (version 4.1.2; NIH) at L4/L5. Fat tissue was defined as −190 to −30 Hounsfield units (HU), lean tissue as 0 to 100 HU, and densities between 0 and −30 HU as undefined. Visceral adipose tissue and abdominal muscle areas were calculated. Bilateral oblique, rectus abdominis, paraspinous, and psoas muscles were segmented within their fascial planes, and muscle area was calculated as pixels with 0 to 100 HU values. Muscle radiodensity, representing fatty infiltration (myosteatosis), was the average HU within a muscle’s fascial plane. Visceral adipose tissue was computed as pixels with −30 to −190 HU in the visceral cavity. Rectus abdominis density, reflecting fat infiltration (lower density indicates greater myosteatosis), was used to assess abdominal myosteatosis due to its proximity to central adipose depots and core stabilizing function. Herein, abdominal myosteatosis is defined by rectus abdominis muscle density.

### 2.3 Primary Outcome

Digit Symbol Coding (DSC), a test of processing speed, and a measure of global cognitive performance and Cognitive Abilities Screening Instrument (CASI, version 2), were obtained in MESA participants at visit 5 (2010–2012) in English, Spanish, Mandarin, and Cantonese. The DSC is a subtest of the Wechsler Adult Intelligence Scale III, with lower scores representing worse performance. For the DSC (range 0–133), participants were presented nine digit–symbol pairs followed by a list of randomly ordered digits, below which they were to write as many corresponding symbols as possible within 120 seconds.^21^

### 2.4 Other variables of interest

Standard questionnaires were used to obtain information on participant demographics, including self-reported age, sex, ethnicity (i.e., non-Hispanic White, Chinese American, African American, Hispanic American) and education (presence of any postsecondary school). Dementia risk factors included diabetes (fasting glucose greater or qual to 126 mg/dL or medication use, as previously described^9^), smoking history, physical activity (by self-report), *APOE* ε4 (*APOE4*) allele status, total serum cholesterol, and systolic blood pressure. These were selected based on the Cardiovascular Risk Factors, Ageing and Dementia risk score.^22^

The CASI (range 0-100; lower score indicates worse performance) includes 25 items representing nine cognitive domains: attention, concentration, orientation, language, verbal fluency, visual construction, abstraction/judgment, and short- and long-term memory. CASI was used as a covariate as a measure of general cognition.

General adiposity was measured as body mass index (BMI) and central adiposity was measured by abdominal visceral fat area from CT scans, as described above.

Levels of cytokines were measured in venous blood collected after a 12-hour fast, processed, and immediately stored at −80 °C. Samples were shipped to the MESA central laboratory (Laboratory for Clinical Biochemistry Research, University of Vermont) for measurement of total cholesterol and cytokines including: Fibroblast Growth Factor (FGF21), Monocyte chemoattractant protein 11(MCP11), C-reactive protein (CRP), adiponectin, insulin, leptin, tumor necrosis factor-alpha (TNF), interleukin-6 (IL6). Concentrations were measured using BioRad Luminex flow cytometry (Millipore).

### 2.5 Statistical Analysis

Descriptive statistics of the full cohort were tabulated and compared across White, Black, Hispanic, and Chinese race/ethnicity/ethnic groups using standard t-tests and chi-square analyses based on variable type. Pairwise correlations were performed between the descriptive characteristics of the population and the main variables (muscle density and DSC) to identify potential confounders. Linear regression models were constructed using muscle density as the primary potential determinant of DSC. In our statistical approach, we carefully considered the relationship of the independent variables to each other to ensure robustness in our regression models. Initially, we included demographic and basic physiological measures such as age, race/ethnicity, sex, muscle area, education level, and general cognition (CASI) to establish a baseline model. To further explore the relationships between abdominal muscle density and DSC, we introduced additional predictors in a phased manner: 1) dementia-related risk factors (such as systolic blood pressure and diabetes) 2) measures of adiposity and 3) levels of circulating cytokines. Each set of variables was added individually to assess their incremental impact on the prediction accuracy and to observe any significant changes in the coefficient of abdominal muscle density. This stepwise approach allowed us to understand the influence of each variable group while managing the correlations among the variables, thus maintaining the clarity and stability of our model estimates. We tested two-way interactions by 1) race/ethnicity, 2) sex and 3) age in the fully adjusted models. All analysis was performed using Stata 17.0 with p<0.05 for two-tailed analyses considered to be significant.

## 3. RESULTS

Characteristics of the full cohort are presented in **Table 1**. Overall, 52% of the participants were women with a mean age 63 ± 9 years with 41% White, 19% Black, 26% Hispanic, and 14% Chinese. Muscle density of individual muscles of the abdomen (oblique, rectus abdominis, paraspinous) and psoas were highly correlated (B=0.69-0.86, p<0.05), (**Supplementary Table 1)**. Ethnic groups differed on all the variables examined (**Supplementary Table 2)**. For example, abdominal muscle density and muscle area were highest in Black adults compared to White, Chinese, and Hispanic groups. The DSC performance was highest among White and Chinese groups, while lower DSC performances were recorded in Black and Hispanic groups. CASI performance was highest in White adults compared to Black, Chinese, and Hispanic groups.

**Table 1:** Population characteristics and correlations or mean differences with Abdominal Myosteatosis measured by Rectus Abdominis Muscle Density and DSC.

|  | Mean (SD) or N (%) in the full cohort. | Correlations or Mean Differences with main variable of interest |  |
| --- | --- | --- | --- |
|  |  | Rectus Abdominis Muscle Density (Hu) | DSC |
| Age (in years) | 63.0 (9.0) | -0.37* | -0.40* |
| Gender |  |  |  |
| Female | 659 (52.0%) | -8.23* | 0.81 |
| Race/ethnicity |  |  |  |
| White | 524 (41.3%) |  |  |
| Black | 247 (19.5%) | 2.61* | -10.34* |
| Chinese | 172 (13.6%) | 1.22 | -2.11 |
| Hispanic | 325 (25.6%) | -0.39 | -16.85* |
| Education |  |  |  |
| Postsecondary | 851 (67.1%) | 3.14* | 16.37* |
| CASI | 87.2 (10.4) | 0.14* | 0.48* |
| Diabetes |  |  |  |
| Present | 126 (9.9%) | -0.75 | -10.06* |
| Systolic Blood Pressure | 122.5 (20.5) | -0.15* | -0.26* |
| Cigarette Smoking |  |  |  |
| Never | 604 (48.0%) |  |  |
| Current | 516 (41.0%) | 2.84* | -0.22 |
| Former | 140 (11.11%) | 1.78* | 0.09 |
| Cholesterol | 190.68 (35.5) | -0.08* | 0.04 |
| ApoE4 |  |  |  |
| Present | 306 (24.1%) | 0.43 | -1.12 |
| Physical Activity (kcal/kg/week) | 5140.2 (4754.4) | 0.18* | 0.02 |
| BMI (kg/m<sup>2</sup>) | 27.8 (4.94) | -0.14* | -0.09* |
| Rectus Abdominis Muscle Density (Hi) | 36.8 (7.5) | 1.00 | 0.21* |
| Abdominal Visceral adiposity (cm<sup>2</sup>) | 141.83 (66.24) | -0.11* | -0.13* |
| Rectus Abdominis Muscle Area | 9.2 (4.2) | 0.68* | 0.11* |

\*p<0.05
DSC, Digit symbol coding
CASI, cognitive abilities screening instrument
Hu, Hounsfield units
SD, standard deviation

There was a significant negative correlation between abdominal myosteatosis and DSC (**Table 1**). Abdominal myosteatosis was significantly and positively correlated with age, female sex, systolic blood pressure, serum cholesterol, BMI, and abdominal visceral fat area. Abdominal myosteatosis was also significantly and negatively correlated with education, CASI, smoking, physical activity, and abdominal muscle area (**Table 1**).

Abdominal myosteatosis was negatively associated (B=−0.247, p=0.001) with DSC and remained independent after adjustment for age, sex, ethnicity, education, and general cognition as measured by CASI (**Table 3**). In multivariable models that accounted for dementia risk factors, greater abdominal myosteatosis continued to be significantly associated with lower DSC scores (**Table 3**). The significant and negative association between abdominal myosteatosis and DSC were not substantially attenuated by additional adjustments for general adiposity with BMI (B=−0.235, p=0.004) or central adiposity with visceral fat area (B=−0.229, p=0.004).

Among the cytokines associated with myosteatosis, IL-6, FGF-21, and fibrinogen were identified as being associated with lower DSC scores (**Table 2**). These cytokines were evaluated as potential confounders of the relationship between abdominal myosteatosis and DSC. Separate adjustments for IL-6, FGF-21, or fibrinogen in the full model did not attenuate the significant association between abdominal myosteatosis and DSC (p<0.05), **Table 3**. Interactions by ethnicity for the relationship between abdominal myosteatosis and DSC were evaluated using the full model (Model 3). Results indicated that these interactions were not statistically significant for Chinese adults (B=0.243, 95% CI −0.086,0.573,p=0.147), Black adults (B=−0.063, 95% CI −0.355,0.229,p=0.672), or Hispanic adults (B=−0.001, 95% CI −0.247,0.246, p=0.996) compared to White adults. Similarly, interactions by sex (B=0.037, (95% CI, −0.220,0.295, p=0.776) and age (B=0.003, 95% CI −0.008,0.140, p=0.625) in the final model were not statistically significant.

**Table 2:** Pairwise Correlations between Muscle density and Cytokines with Digit Symbol Coding, in Full Cohort.

| **Table 2:** Pairwise Correlations between Muscle density and Cytokines with Digit Symbol Coding, in Full Cohort (N=1268). |  |  |  |  |  |  |  |  |  |
| --- | --- | --- | --- | --- | --- | --- | --- | --- | --- |
|  | IL-6 | FGF-21 | Insulin | Leptin | Fibrinogen | CRP | MCP11 | ADPN | TNFa |
| Rectus Abdominis Muscle Density | -0.094** | -0.143* | -0.026 | -0.353** | -0.240** | -0.172 | -0.135 | -0.315** | -0.003 |
| DSC | -0.102** | -0.073* | -0.072* | -0.041 | -0.168** | 0.059* | 0.031 | 0.003 | -0.048 |

\*p<0.05
\*\*p<.01
DSC, Digit symbol coding
IL-6, Interleukin 6
FGF-21, Fibroblast Growth Factor 21
MCP11: Monocyte chemoattractant protein 11
TNFa: Tumor necrosis factor alpha
AMD: Abdominal Muscle density
ADPN: Adiponectin

**TABLE 3.**
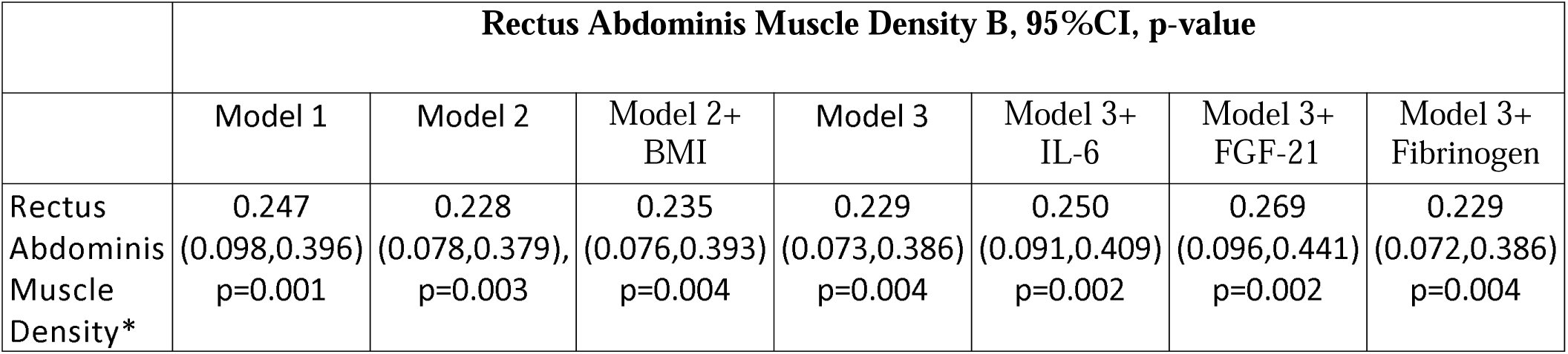
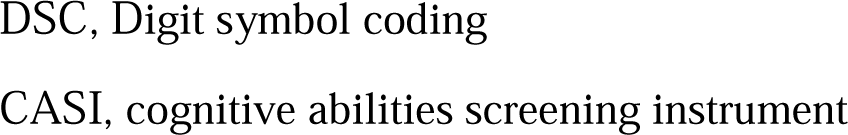
Beta coefficients with 95% CI of multivariable regression models with rectus abdominis muscle and DSC (outcome). All models are adjusted for age, race/ethnicity, gender, education, and CASI. Model 2 is Model 1+ adjusted for physical activity, APOe4 allele status, presence of diabetes, serum cholesterol, smoking, systolic blood pressure. Model 3 is Model 2+ Visceral Fat.

## 4. Discussion

In this multiethnic sample of adults, greater abdominal myosteatosis was associated with lower processing speed scores. This finding was independent of ethnicity, sex, multiple dementia risk factors, central and general adiposity, and levels of pro-inflammatory cytokines. Collectively, these findings suggest that abdominal myosteatosis may offer unique clinical insight as a diagnostic tool and a novel rehabilitation target for processing speed performance in diverse populations of middle aged and older adults.

Recent investigations have reported thigh inter-muscular adipose tissue and calf muscle density to be associated with mini-mental status decline and processing speed in White and Black older Americans and African Caribbean women, respectively.^11,12^ Moreover, studies evaluating association of myosteatosis with markers of brain health on MRI were conducted in homogenous populations of European adults.^14,23^ Our study expands on previous research by including a diverse, multiethnic cohort of White, Chinese, Black, and Hispanic U.S. adults, analyzing interactions by ethnicity and utilizing abdominal myosteatosis as a marker, rather than leg myosteatosis.

Our finding that abdominal myosteatosis remained associated to processing speed performance even after adjusting for dementia-related risk factors is supported by the literature. When we adjusted for established dementia risk factors (i.e., education, diabetes, systolic blood pressure, smoking, physical activity, serum cholesterol, and ApoE4 status), only education, systolic blood pressure, and smoking were significantly associated with the presence of abdominal myosteatosis. Collectively, these results suggest that myosteatosis may be independently associated with processing speed and the link between myosteatosis and cognition health may be due to other factors beyond established dementia risk factors.

The positive correlation between myosteatosis and pro-inflammatory cytokines has been previously reported in this cohort.^8^ Contrary to our hypotheses, levels of pro-inflammatory cytokines did not modify the association between abdominal myosteatosis and processing speed. However, this finding is in line with our recent investigation reporting that leptin, adiponectin, and IL-6 do not considerably change the association of thigh myosteatosis with cognitive decline.^11^ In the current study, we observed a significant bivariate relationship of abdominal myosteatosis to the pro-inflammatory cytokines of IL-6, FGF-21, and fibrinogen, as well as a relationship of abdominal myosteatosis to processing speed.^8^ Our findings build upon the existing literature and suggest that pro-inflammatory serum cytokines do not considerably change the association between myosteatosis and cognitive health.^11^

Interestingly, muscle area, visceral fat, and BMI measures were not significantly associated with processing speed in models including abdominal myosteatosis. This is similar to the findings in an independent cohort, where myosteatosis was associated with cognitive decline and muscle area, central adiposity, and overall adiposity were not.^11,12^ In the current study, visceral fat area and BMI were not significant in multivariable models in the presence of abdominal myosteatosis despite visceral fat and BMI having statistically significant negative bivariate correlations with DSC. This may support the possibility that muscle fat depots and visceral adiposity may share some similar mechanisms in their impact on cognition, perhaps via similar effects on cardiometabolic health. Cardiometabolic health is one biologically plausible mechanism as it is associated to cerebral white matter hyperintensity volume, a direct measure of cerebrovascular injury of the brain and cognitive processing speed performance.^31^ Lastly, this highlights a direction for future research to evaluate the contributing mechanisms, including cardiometabolic health, for the relationship between skeletal myosteatosis and processing speed.

Our study has limitations. First, the various measurements were single measures that were not obtained concurrently; cytokines were measured 1 year prior to abdominal muscle density, while cognition was measured 7 years after. While these measures change slowly, using repeated and concurrent measures would provide better estimates of these relationships and, in particular, effects on cognitive change and would decrease temporal bias. Prior work has demonstrated that increases in myosteatosis were associated with cognitive decline over five years.^11^ It is possible selection bias may have occurred between clinical exams 2 and 3 and clinical exam 5. To assess the potential impact of this bias, we analyzed the baseline characteristics of participants who were evaluated for myosteatosis at exams 2 and 3 but did not have cognitive data at exam 5. These characteristics are detailed in **Supplementary Table 2** for included participants and **Supplementary Table 3** for those excluded from the study. Comparative analysis revealed no significant differences between the groups in terms of smoking status, ApoE4 allele presence, serum cholesterol levels, physical activity, and BMI. However, the excluded participants were slightly older, more likely to be male, and had lower levels of postsecondary education (60.4% versus 67.1%). Additionally, there were slight increases in diabetes prevalence and notably higher systolic blood pressure rates (130 versus 122) among those excluded. The exclusion of these participants from the final analysis may have introduced a bias, making it unclear how their absence definitively affected our results. Therefore, additional studies are needed. Furthermore, our sample size may have limited the power to detect interactions based on sex, ethnicity, and measures of central adiposity within the analysis of abdominal myosteatosis and DSC. Lastly, except for IL-6, which is considered both adipokine and myokine, we were limited to investigating relevant cytokines previously collected in MESA. Even though these cytokines are known to be involved in the hypothesized mechanisms by which muscle health may impact cognition: inflammation, cardio-metabolic health, and muscle-brain crosstalk, future studies are needed to further investigate other relevant myokines and mediators to better understand a potential causal relationship between muscle and cognition.

These limitations notwithstanding, our findings may have biologic plausibility and clinical implications. There is increasing recognition of the bidirectional crosstalk between muscle and the brain, highlighting the emerging role of muscle as a secretory organ. Muscle’s diverse secretory profile, which includes myokines such as nucleic acids, small molecules, peptides, and hormones, either freely circulating or contained within extracellular vesicles, presents intriguing mechanisms for muscle-to-brain interplay.^5^ For example, myostatin, a myokine, has been reported to be negatively associated with circulating amyloid burden.^24^ Future research should further explore the various secretory mediators released by muscle and their potential roles in mediating muscle’s relationship to processing speed. From a clinical perspective, the widespread availability of abdominal CT scans offers a potential opportunity to use abdominal myosteatosis as a diagnostic biomarker, as well as a potentially modifiable therapeutic target. While physical activity interventions are strongly associated with improvements in cognitive and mental health, they have largely focused on aerobic exercise.^25,26^ It is worth considering that interventions targeting improvements in muscle density, which is closely correlated with strength and power, may also hold promise. Thus, strength-training and muscle intervention protocols designed to enhance muscle density deserve future consideration as potential strategies for improving cognitive outcomes.

## 5. Conclusion

Our collective findings suggest that abdominal myosteatosis is negatively associated with cognitive processing speed in a multiethnic cohort and is independent of traditional dementia risk factors. Moving forward, research should focus on investigating the impact of myosteatosis on cardiometabolic health and muscle brain crosstalk. Given the known association between physical function and cognitive health in aging adults, it is reasonable to think that myosteatosis may offer new insight into physical function’s protective influence on cognition.

## Funding

This research was supported by contracts 75N92020D00001, HHSN268201500003I, N01-HC-95159, 75N92020D00005, N01-HC-95160, 75N92020D00002, N01-HC-95161, 75N92020D00003, N01-HC-95162, 75N92020D00006, N01-HC-95163, 75N92020D00004, N01-HC-95164, 75N92020D00007, N01-HC-95165, N01-HC-95166, N01-HC-95167, N01-HC-95168 and N01-HC-95169 from the National Heart, Lung, and Blood Institute, and by grants UL1-TR-000040, UL1-TR-001079, and UL1-TR-001420 from the National Center for Advancing Translational Sciences (NCATS). The authors thank the other investigators, the staff, and the participants of the MESA study for their valuable contributions. A full list of participating MESA investigators and institutions can be found at http://www.mesa-nhlbi.org.

**This paper has been reviewed and approved by the MESA Publications and Presentations Committee.**

## Contributions of Authors

BLM drafted the manuscript and revised analyses.

CR and IM developed analysis plan, completed analyses, and revised the manuscript. ET completed analyses. BLM, IM, MAA, TH, IN, ET, CR all critically reviewed and edited the manuscript and MAA secured funding for this project and supervised the collection of body composition data.

## Data Availability

Data from MESA are available to investigators.

---

**Supplementary Table 1:** Pairwise Pearson Correlations of Muscle Density between muscle groups.

|  | Psoas | Oblique | Paraspinous |
| --- | --- | --- | --- |
| Oblique | 0.69, p<.0001 |  |  |
| Paraspinous | 0.77, p<.0001 | 0.77, p<.0001 |  |
| Rectus Abdominis | 0.52, p<.0001 | 0.77, p<.0001 | 0.67, p<.0001 |

**Supplementary Table 2.** Characteristics of participants with muscle density data at Exam 2 and Cognitive Outcomes at Exam 5 by Race/ethnicity/Ethnicity. Mean (SD) or n (%).

|  | All<br>N = 1268 | Race/ethnicity / Ethnicity |  |  |  | P-value |
| --- | --- | --- | --- | --- | --- | --- |
|  |  | White<br>N = 524 | Black<br>N=247 | Chinese<br>N = 172 | Hispanic<br>N = 325 |  |
| Age (in years) | 63.0 (9.0) | 64.0 (98.99) | 62.0 (9.21) | 62.3 (8.82) | 61.9 (8.82) | 0.0058 |
| Sex<br>Female | 659 (51.97%) | 262 (50.0%) | 146 (59.11%) | 82 (47.67%) | 169 (52.00%) | 0.0679 |
| Education<br>Postsecondary | 851 (67.11%) | 426 (81.30%) | 182 (73.68%) | 112 (65.12%) | 131 (40.31%) | <.0001 |
| Diabetes<br>Present | 126 (9.94%) | 32 (6.11%) | 33 (13.36%) | 18 (10.47%) | 43 (13.23%) | 0.0012 |
| Systolic Blood<br>Pressure (mm Hg) | 122.5 (20.5) | 120.5 (19.3) | 129.6 (22.1) | 118.7 (18.8) | 122.4 (20.7) | <.0001 |
| Cigarette Smoking<br>Never<br>Former<br>Current | 604 (47.94%)<br>516 (40.95%)<br>140 (11.11%) | 210 (40.54%)<br>246 (47.49%)<br>62 (11.97%) | 105 (42.68%)<br>103 (41.87%)<br>38 (15.45%) | 126 (73.26%)<br>41 (23.84%)<br>5 (2.91%) | 163 (50.31%)<br>126 (38.89%)<br>35 (10.80%) | <.0001 |
| ApoE4<br>Present | 306 (24.13%) | 125 (23.85%) | 86 (34.82%) | 25 (14.53%) | 70 (21.54%) | <.0001 |
| Cholesterol (mg/dL) | 190.68 (35.48) | 188.16 (34.58) | 190.96 (35.71) | 188.65 (30.90) | 195.61 (38.51) | 0.0327 |
| Physical Activity<br>(kcal/kg/week) | 5140.18<br>(4754.42) | 4958.02<br>(3903.71) | 5673.18<br>(5441.66) | 3884.29<br>(4532.46) | 5693.46<br>(5390.39) | <.0001 |
| BMI (kg/m<sup>2</sup>) | 27.82 (4.94) | 27.57 (4.77) | 29.16 (5.05) | 24.31 (3.26) | 29.05 (4.93) | <.0001 |
| Visceral Fat Area (cm<sup>2</sup>) | 141.83 (66.24) | 154.20 (72.37) | 122.10 (56.41) | 113.18 (50.31) | 152.06 (62.16) | <.0001 |
| Rectus Abdominis Muscle Area (cm<sup>2</sup>) | 9.19 (4.18) | 9.22 (4.02) | 10.14 (4.80) | 7.69 (3.16) | 9.22 (4.17) | <.0001 |
| Rectus Abdominis Muscle density (Hu) | 36.87.37 (7.50) | 36.30 (7.31) | 38.88 (7.07) | 37.52 (7.13) | 35.91 (8.02) | <.0001 |
| DSC | 50.15 (18.56) | 56.77 (15.87) | 46.43 (16.96) | 54.66 (18.26) | 39.92 (18.69) | <.0001 |
| CASI | 87.18 (10.36) | 91.12 (8.58) | 86.11 (9.53) | 84.71 (12.02) | 82.94 (10.40) | <.0001 |

DSC, Digit symbol coding
CASI, cognitive abilities screening instrument

**Supplementary Table 3:** Demographic Data of participants who had myosteatosis measurements but cognitive testing at Exam 6 was missing.

|  | All<br>N = 490 | Race/ethnicity / Ethnicity |  |  |  |
| --- | --- | --- | --- | --- | --- |
|  |  | Black<br>N = 120 | Chinese<br>N=75 | Hispanic / Latino<br>N = 111 | White<br>N = 184 |
| Age (in years) | 65.8 (10.4) | 65.8 (10.2) | 68.1 (10.6) | 64.1 (10.5) | 65.9 (10.2) |
| Sex |  |  |  |  |  |
| Female | 239 (48.8%) | 64 (53.3%) | 36 (48.0%) | 54 (48.7%) | 85 (46.2%) |
| Education |  |  |  |  |  |
| Postsecondary | 296 (60.4%) | 84 (70.0%) | 39 (52.0%) | 30 (27.0%) | 143 (77.7%) |
| Diabetes |  |  |  |  |  |
| Present | 39 (7.96%) | 14 (11.7%) | 6 (8.0%) | 9 (8.1%) | 10 (5.4%) |
| SBP | 130.8 (23.4) | 134.4 (22.8) | 131.8 (24.1) | 132.2 (26.2) | 127.3 (21.2) |
| Cigarette Smoking |  |  |  |  |  |
| Current | 64 (13.1%) | 20 (16.7%) | 6 (8.1%) | 15 (13.5%) | 23 (12.6%) |
| Former | 180 (36.9%) | 46 (38.3%) | 15 (20.3%) | 37 (33.3%) | 82 (44.8%) |
| Never | 244 (50.0%) | 54 (45.0%) | 53 (71.6%) | 59 (53.2%) | 78 (42.6%) |
| ApoE4 |  |  |  |  |  |
| Present | 117 (23.9%) | 36 (30.0%) | 10 (13.3%) | 26 (23.4%) | 45 (24.5%) |
| Cholesterol (mg/dL) | 194.3 (32.1) | 190.0 (36.1) | 192.3 (30.0) | 196.8 (30.2) | 196.4 (31.0) |
| Physical Activity<br>(kcal/kg/week) | 5310.4 (5360.9) | 5894.8 (6245.4) | 3440.9 (3145.6) | 5507.5 (6445.2) | 5573.7 (4546.3) |
| BMI (kg/m2) | 27.3 (5.0) | 29.3 (5.8) | 23.6 (2.9) | 28.5 (4.6) | 26.9 (4.5) |
| Rectus Abdominis<br>Muscle Density | 34.3 (8.4) | 36.2 (8.6) | 32.7 (9.1) | 33.2 (7.8) | 34.2 (8.1) |

